# Prevalence of antibodies to SARS-CoV-2 in healthy blood donors in New York

**DOI:** 10.1101/2020.10.19.20215368

**Authors:** Kathy Kamath, Elisabeth Baum-Jones, Gregory Jordan, Winston Haynes, Rebecca Waitz, John Shon, Steve Kujawa, Lyn Fitzgibbons, Debra Kessler, Larry Luchsinger, Yale IMPACT Team, Patrick Daugherty

## Abstract

Despite the high level of morbidity and mortality worldwide, there is increasing evidence for asymptomatic carriers of the novel coronavirus SARS-CoV-2. We analyzed blood specimens from 1,559 healthy blood donors, collected in the greater New York metropolitan area between the months of March and July 2020 for antibodies to SARS-CoV-2 virus. Using our proprietary technology, SERA (Serum Epitope Repertoire Analysis), we observed a significant increase in SARS-CoV-2 seropositivity rates over the four-month period, from 0% [95% CI: 0 - 1.5%] (March) to 11.6% [6.0 - 21.2%] (July). Follow-up ELISA tests using S1 and nucleocapsid viral proteins confirmed most of these results. Our findings are consistent with seroprevalence studies within the region and with reports that SARS-COV-2 infections can be asymptomatic or cause only mild symptoms.

**IMPORTANCE:** The COVID-19 pandemic, caused by the novel coronavirus SARS-CoV-2, has caused vast morbidity and mortality worldwide, yet several studies indicate that there may be a significant number of infected people who are asymptomatic or exhibit mild symptoms. In this study, samples were collected from healthy blood donors in a region of rapidly increasing disease burden (New York metropolitan area) and we hypothesized that a subset would be seropositive to SARS-CoV-2. People who experienced mild or no symptoms during SARS-CoV-2 infection may represent a source for convalescent plasma donors.

## OBSERVATION

The COVID-19 pandemic, caused by the novel coronavirus SARS-CoV-2, has exerted high morbidity worldwide, with mortality rates reported greater than 10% for some age groups [1]. Yet several studies have indicated that there may be a significant number of people who are asymptomatic or exhibit mild symptoms [2 - 4], including in the New York metropolitan region [5, 6]. We developed an assay to detect IgG and IgM antibodies to SARS-CoV-2 virus and analyzed plasma samples from healthy blood donors collected during the early months of the pandemic in that region.

### Development of SERA assay for antibodies to SARS-CoV-2

Our technology, Serum Epitope Repertoire Analysis (SERA) enables discovery and semi-quantitative detection of antibody epitopes with high resolution that can be mapped to eliciting antigens and organisms. The SERA assay platform has been described in detail elsewhere [7], but briefly, it utilizes a random bacterial display 12-mer peptide library of 10^10^ diversity in conjunction with NGS to collect and sequence the set of peptides that represent an individual’s serological antibody epitope repertoire. The shared epitopes targeted by antibody repertoires are then identified by custom bioinformatics algorithms [8], using cohort-based subtractive analysis of the mimotope repertoires. For the development of SERA panels to detect IgG and IgM antibodies against SARS-CoV-2, serum samples from confirmed COVID-19 patients (positive for SARS-CoV-2 nucleic assay testing (NAT)) were acquired from the Yale IMPACT biorepository and the Santa Barbara Cottage Hospital (Santa Barbara, CA) in accordance with BRISQ guidelines [9] and the SBCH IRB. The majority of subjects were hospitalized and serum samples were collected between 0 and 57 days post symptom onset. For the discovery of SARS-CoV-2 IgG and IgM antibody epitope motifs, we applied SERA to a Discovery cohort of 164 COVID-19 samples, and 430 (for IgM panel) and 497 (for IgG panel) pre-pandemic healthy controls (**Table S1**). The resulting epitope motifs were compiled into IgG and IgM panels, and the enrichment of each panel motif was normalized and summed to generate a composite diagnostic score [7]. A seropositivity cut-off value of 25 was established to yield a specificity of > 99% for either panel. In an independent verification set of 250 COVID-19 specimens for which we had days since symptom onset data, the sensitivity of SERA IgG and IgM at >10 days post symptom onset was ≥ 91% (**Figure 1**). In a cohort of 1,500 pre-pandemic samples, the specificity of the SERA SARS-CoV-2 IgG panel was 99.3% and the IgM panel was 99.1%; the specificity of the IgG and IgM panels together was 98.7%.

**Figure 1:**
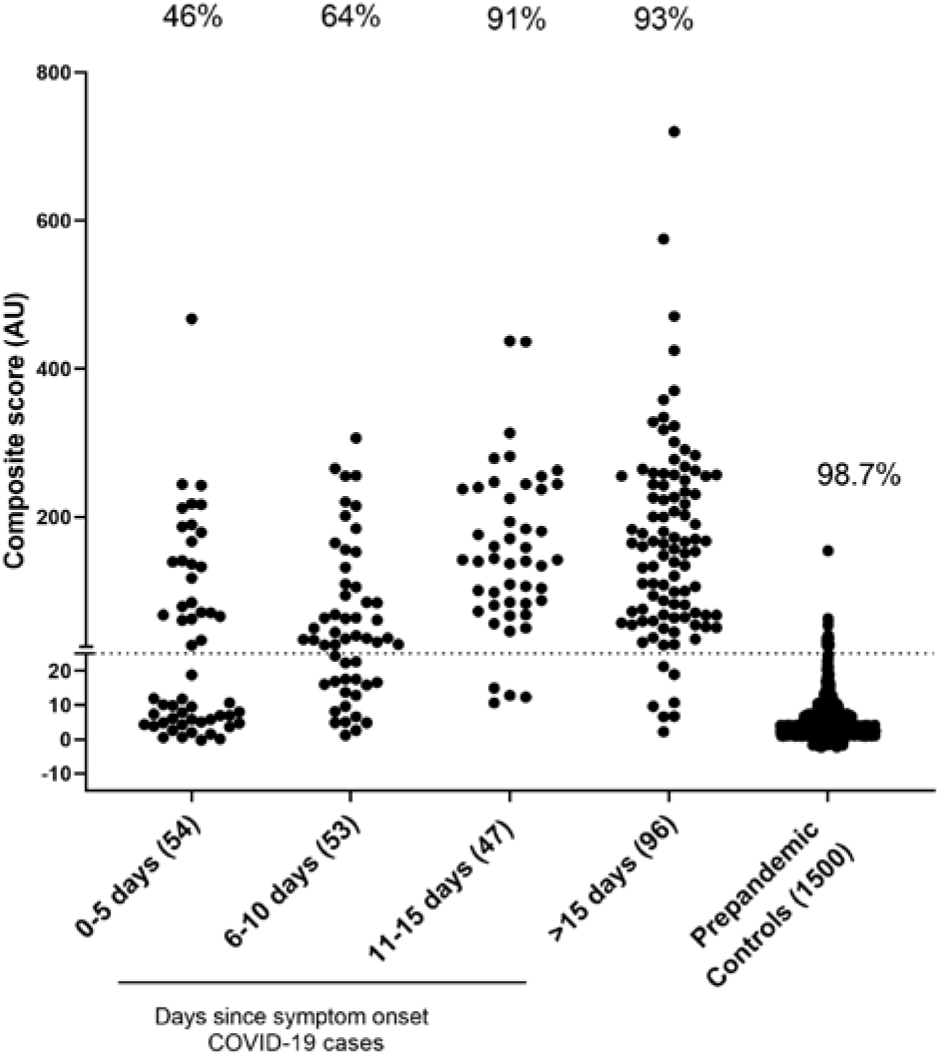
Performance SERA assay to detect IgG and IgM antibodies to SARS-CoV-2. SERA IgG+IgM performance on a validation cohort of SARS-CoV-2 NAT+ patients. The maximum SERA score for IgG or IgM is shown. Samples above a cutoff of 25 are classified as seropositive. COVID-19 cases are indicated by days post symptom onset. Panel sensitivity at the specified time intervals are shown above each group. Specificity on a cohort of 1,500 pre-pandemic controls was 99.3% for IgG, 99.1% for IgM, 98.7% for IgG or IgM, and 100% if a case was positive for IgG and IgM (no pre-pandemic validation controls were positive for both IgG and IgM concomitantly).

### Sample collection from healthy blood donors

We initiated a collaboration with Blood Centers of America to collect plasma from healthy donors throughout the US with a primary objective of increasing our healthy subject cohort. Collection and screening began in New York at the New York Blood Center (NYBC) in March 2020. Individuals who were suspected of having COVID-19 were deferred from donation for 14 days after resolution of symptoms. For this study, a total of 1,559 whole blood samples from healthy blood donors were sent to our laboratory within 2 weeks of collection, in seven shipments. The demographics of these samples are shown in **Table S2**. Upon arrival at our laboratory, blood plasma was isolated, screened by SERA assay and the results were uploaded to the epitope repertoire database.

### Prevalence of antibodies to SARS-CoV-2 in blood donors

For each sample analyzed using the SERA assay, the epitope repertoires are compiled into a database that enables assessment of any previously or future developed disease-specific SERA panel, without the need to re-test the sample. We applied the SARS-CoV-2 IgG panel retrospectively to samples collected during the period March 1 through July 7, 2020. Of 1,559 blood samples processed, a total of 68 had IgG antibodies against SARS-CoV-2 on the SERA assay. As collection proceeded, the seropositive rate increased from 0% [95% CI: 0 - 1.5%] to 11.6% [6.0 - 21.2%] (**Figure 2, Table S3**). Eighty-four percent (57/68) of these samples were also positive for IgG antibodies to SARS-CoV-2 Spike S1 and nucleocapsid proteins by follow-up ELISAs (**Supplemental Text 1**), providing independent verification of the presence of a SARS-CoV-2 antibody response in these subjects.

**Figure 2.**
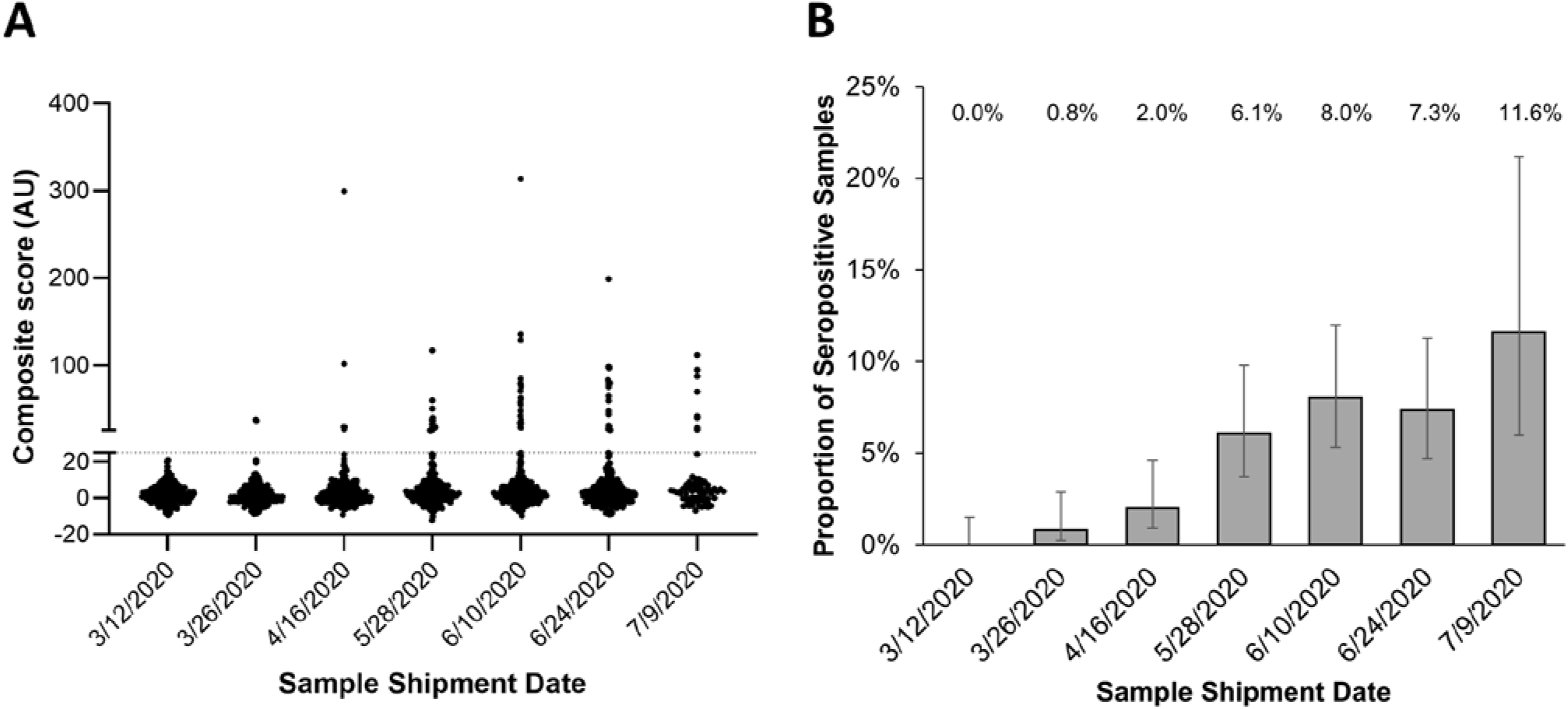
Seropositivity rate of SARS-CoV-2 based on SERA IgG increases over time in healthy, asymptomatic blood donors. A) SERA SARS-CoV-2 IgG scores and B) percentage of samples that were SARS-CoV-2 IgG positive from healthy blood donors by shipment date. Error bars in (B) represent 95% CI using Wilson score. Shipment dates are shown on the x-axis; blood samples were collected within 2 weeks prior to shipment. Each shipment included 245-250 subjects except the final shipment (n = 69).

Because the age of donors were not equally distributed amongst the sample shipments, we interrogated whether age or shipment date was a stronger predictor of the seropositivity observed. GLM regression analysis of SARS-CoV-2 seropositivity relative to donor age or shipment date revealed that sample shipment date contributed more significantly (p=5.5e-8) to the prediction of seropositivity than age (p=0.35) (**Supplemental Figure 1**).

### Concluding remarks

This study provides insight into trends of rate of seropositivity to SARS-CoV-2 among asymptomatic blood donors in the metropolitan New York area during the early months of the COVID-19 pandemic but was not designed to measure population-wide seroprevalence. Nonetheless, our results were consistent with reports that a significant number of SARS-CoV-2 infected people in this region may be asymptomatic or present with mild symptoms [5, 6].

Here we demonstrated the utility of SERA for serosurveillance studies of emerging infections through retrospective analysis of archived data in populations. Previously collected data was simply re-analyzed for SARS-CoV-2 associated epitopes, thus mitigating the need to store samples or perform additional assays in a surveillance setting. In addition, the new panel was applied retrospectively *in silico* to large pre-pandemic cohorts in panel development and verification phases, demonstrating high specificity without the need to perform additional immunoassays. Finally, applying the panel to cohorts afforded the opportunity to identify subjects with presumptive asymptomatic or mild disease for research purposes amongst the blood donor population from New York and surrounding areas.

Our results raise the question of whether a larger number of convalescent plasma donors could be identified among healthy, asymptomatic donors. However, there is concern about providing antibody results to blood donors without an improved understanding of their immunological significance. We also demonstrate the utility of SERA for monitoring the antibody response to emerging infections.

## Data Availability

Data from SERA assay is not available.

## SUPPLEMENTAL MATERIALS

**Supplemental Text 1: Spike and Nucleocapsid protein ELISA**. For confirmatory ELISA testing, 50 µL of SARS-CoV-2 Spike S1 or nucleocapsid recombinant proteins (ACROBiosystems cat# S1N-C52H3 and NUN-C5227) were coated on ELISA plates at 0.5 µg/mL. Plates were blocked with 5% nonfat dry milk (NFDM) in PBS followed by incubation with plasma diluted 1:250 in PBS. Wells were washed 3X with PBS with 0.1% tween20 (PBST) and incubated with anti-human IgG conjugated to HRP (Jackson Immuno Research labs cat# 109-035-088) at 1:10,000. Wells were washed 4X with PBST and developed with TMB solution. Cut-off values for ELISAs were established based on 400 pre-pandemic controls.

**Supplemental Figure S1:**
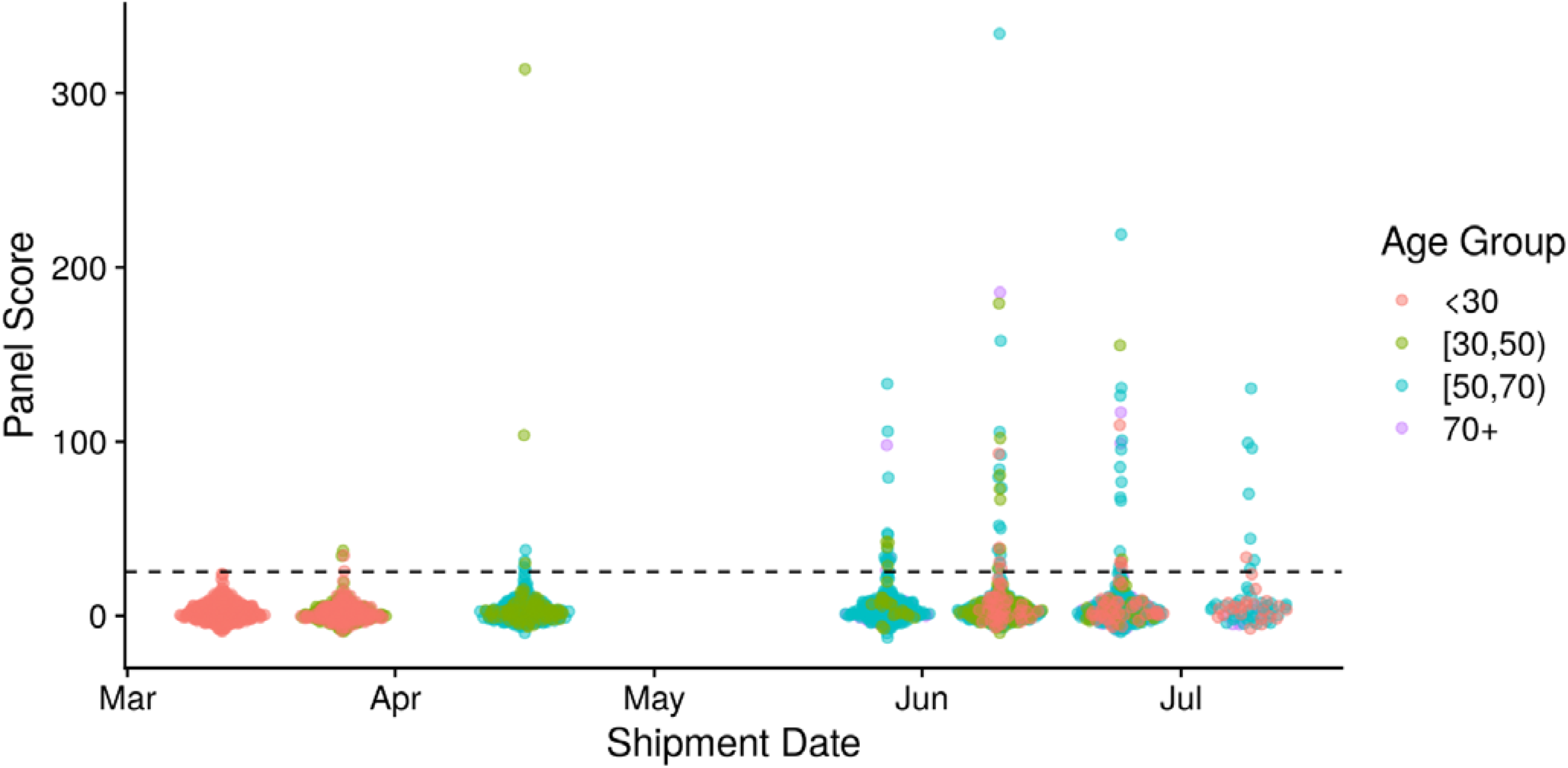
SARS-CoV-2 seropositivity vs. shipment date for each specimen. GLM regression analysis of seropositivity to SARS-CoV-2 relative to the age of the patient and the shipment date of the sample. Shipment date contributed more significantly (p=5.5e-8) to the prediction of seropositivity than age (p=0.35). When we performed the same regression for SARS-CoV-2 seropositivity against age alone, the age coefficient was significant (p=5.5e-5). Similarly, regression for SARS-CoV-2 seropositivity against shipment date alone yielded a highly significant shipment date coefficient (p=1.0e-9). These results suggest that shipment date, not the age bias, is the dominant factor contributing to the increased positivity rates.

**Table S1:** Sample cohorts for discovery and verification of SARS-CoV-2 epitope motif panels.

|  | Sample Cohort | COVID-19 Disease Severity |  |  |  | Total n |
| --- | --- | --- | --- | --- | --- | --- |
|  |  | Asymp | Mild/<br>Moderate | Severe | Moderate<br>or Severe* |  |
| Discovery Set | Confirmed COVID-19 (Yale IMPACT) | 5 | 103 | 45 | 11 | 164 |
|  | Pre-pandemic controls | - | - | - | - | 430 (IgM) or 497 (IgG) |
| Verification Set | Confirmed COVID-19 (Yale IMPACT) | 8 | 20 | 17 | 143 | 191 |
|  | Confirmed COVID-19 (SBCH) | 4 | 54 | 22 | - | 81 |
|  | Mayo Clinic pre-pandemic controls | - | - | - | - | 1,500 |

\*Subjects were hospitalized but intubation status was not known

**Table S2:** Demographics of Healthy Blood Donors.

| Donor characteristics |  | n | % |
| --- | --- | --- | --- |
| **Sex** | Female | 755 | 48 |
|  | Male | 804 | 52 |
| **Age** | 17-25 | 395 | 25 |
|  | 26-35 | 200 | 13 |
|  | 36-45 | 164 | 11 |
|  | 46-55 | 206 | 13 |
|  | 56-65 | 370 | 24 |
|  | 66-80 | 224 | 14 |
| **Race** | White | 396 | 25 |
|  | Black | 350 | 22 |
|  | Hispanic/Latino | 454 | 29 |
|  | Asian/Pacific Islander | 358 | 23 |
|  | Other | 1 | 0.1 |

**Table S3.**
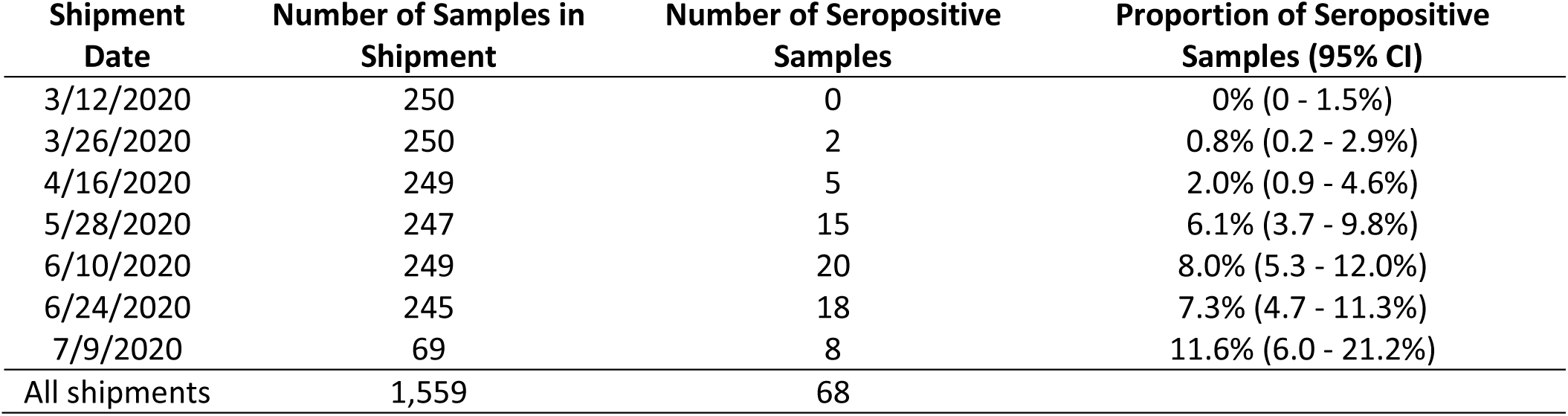
Blood donor samples analyzed and prevalence of seropositive samples per shipment.

